# Rare germline disorders implicate long non-coding RNAs disrupted by chromosomal structural rearrangements

**DOI:** 10.1101/2024.06.16.24307499

**Authors:** Rebecca E. Andersen, Ibrahim F. Alkuraya, Abna Ajeesh, Tyler Sakamoto, Elijah L. Mena, Sami S. Amr, Hila Romi, Margaret A. Kenna, Caroline D. Robson, Ellen S. Wilch, Katarena Nalbandian, Raul Piña-Aguilar, Christopher A. Walsh, Cynthia C. Morton

**Affiliations:** Division of Genetics and Genomics and Manton Center for Orphan Diseases, Boston Children’s Hospital, Boston, MA, USA; Harvard Medical School, Boston, MA, USA; Broad Institute of MIT and Harvard, Cambridge, MA, USA; Department of Obstetrics and Gynecology, Brigham and Women’s Hospital, Boston, MA, USA; Harvard College, Cambridge, MA, USA; Division of Genetics, Department of Genetics, Brigham and Women’s Hospital, Boston, MA, USA; Department of Pathology, Brigham and Women’s Hospital, Boston, MA, USA; Department of Otolaryngology, Boston Children’s Hospital, Boston, MA, USA; Department of Radiology, Boston Children’s Hospital, Boston, MA, USA; Howard Hughes Medical Institute, Boston, MA, USA; University of Manchester, Manchester Center for Audiology and Deafness, UK

## Abstract

In recent years, there has been increased focus on exploring the role the non-protein-coding genome plays in Mendelian disorders. One class of particular interest is long non-coding RNAs (lncRNAs), which has recently been implicated in the regulation of diverse molecular processes. However, because lncRNAs do not encode protein, there is uncertainty regarding what constitutes a pathogenic lncRNA variant, and thus annotating such elements is challenging. The Developmental Genome Anatomy Project (DGAP) and similar projects recruit individuals with apparently balanced chromosomal abnormalities (BCAs) that disrupt or dysregulate genes in order to annotate the human genome. We hypothesized that rearrangements disrupting lncRNAs could be the underlying genetic etiology for the phenotypes of a subset of these individuals. Thus, we assessed 279 cases with BCAs and selected 191 cases with simple BCAs (breakpoints at only two genomic locations) for further analysis of lncRNA disruptions. From these, we identified 66 cases in which the chromosomal rearrangements directly disrupt lncRNAs. Strikingly, the lncRNAs *MEF2C-AS1* and *ENSG00000257522* are each disrupted in two unrelated cases. Furthermore, in 30 cases, no genes of any other class aside from lncRNAs are directly disrupted, consistent with the hypothesis that lncRNA disruptions could underly the phenotypes of these individuals. To showcase the power of this genomic approach for annotating lncRNAs, here we focus on clinical reports and genetic analysis of two individuals with BCAs and additionally highlight six individuals with likely developmental etiologies due to lncRNA disruptions.

## Introduction

Only ∼2% of the human genome directly codes for proteins. Among the approximately 20,000 protein-coding genes, 4,000 have been implicated in Mendelian diseases (Avraham et al. 2022). The non-protein coding genome comprises a diverse array of elements including those that transcribe long non-coding RNA molecules (lncRNAs). lncRNAs are transcripts of at least 200 nucleotides in length but are not translated into proteins (Kopp and Mendell 2018). Current transcriptome annotations (Frankish et al. 2021) suggest that there are nearly 20,000 human lncRNAs, and the expression of many of these lncRNAs is highly regulated. However, the biological roles of most lncRNAs remain to be determined, which is made challenging by the fact that lncRNAs can have such a wide variety of different functions. Some lncRNAs regulate the transcription of nearby genes, whereas others regulate other biological processes, including splicing and translation (Taniue and Akimitsu 2021; Statello et al. 2021). One particularly intriguing class of lncRNAs is that of divergent lncRNAs, which have transcriptional start sites (TSSs) within 5kb of another gene and are transcribed in the opposite direction in a “head-to-head” configuration. Divergent lncRNAs have generally been associated with regulation of their neighboring gene, particularly when the neighbor is a transcription factor (Luo et al. 2016; Wang et al. 2020). More broadly, lncRNAs of several classes have been found to affect the expression of their neighboring genes (Statello et al. 2021), providing a mechanism for such lncRNAs to modulate key biological processes.

As a testament to the importance of lncRNAs in normal development, deletions of certain lncRNAs have led to lethal phenotypes in mice as well as abnormal development of the neocortex, lung, gastrointestinal tract, and heart (Sauvageau et al. 2013; Feyder and Goff 2016; Mattick et al. 2023). In addition, lncRNAs have been implicated in cancer and shown to affect cell division, metabolism, and tumor-host interactions. Thus, lncRNAs are essential to maintaining proper cellular homeostasis (Statello et al. 2021). A prior observation of a variant in a lncRNA causing human disease in a Mendelian fashion is a 27 – 63 kb deletion of a locus that encompasses a lncRNA upstream of the engrailed-1 gene (*EN1*), which resulted in congenital limb abnormalities even though *EN1* itself was not disrupted (Allou et al. 2021). Most recently, a pre-print manuscript (Ganesh et al. 2024) reported three individuals with deletions in *CHASERR*, a lncRNA proximal to *CHD2*, a protein-coding gene that causes developmental and epileptic encephalopathy. Intriguingly, disruption of *CHASERR* leads to increased expression of *CHD2* in *cis*, leading to a distinct clinical presentation compared to individuals with *CHD2* haploinsufficiency.

Given the importance of lncRNAs to gaining a better understanding of developmental biology and improving clinical diagnoses, it is important to develop a better functional assessment of lncRNAs in the human genome. However, the methods of annotating protein-coding genes are not typically applicable to non-coding genes due to their fundamental differences (Mattick et al. 2023). For instance, single nucleotide variants (SNVs) can result in nonsense mutations that prematurely terminate proteins, and indels can cause translational frame shifts that alter the entire downstream amino acid sequence of a protein. However, because lncRNAs do not encode proteins, it is unclear what affect if any such mutations may have on lncRNAs. The study of lncRNAs remains to be elucidated with regard to definitive consensus over what constitutes a pathogenic lncRNA variant.

A prior landmark development in expanding a focus on DNA beyond protein-coding genes to the 3D genome was the discovery of topologically associated domains (TADs). TADs are megabase-sized genomic segments partitioning the genome into large regulatory units with frequent intra-domain chromatin interactions but relatively rare inter-domain interactions (Lupiáñez et al. 2015). Conserved across different cell types and species, they are considered crucial for spatiotemporal gene expression patterns. Topological boundary regions (TBRs) block interactions between adjacent TADs, and TBR disruption by chromosomal structural rearrangements can result in rewiring of genomic regulators leading to abnormal clinical phenotypes. Rigorous interpretation of clinical phenotypes requires assessment of the boundaries of TADs following a chromosomal structural rearrangement because complex phenotypes may be dissected from rearrangements that reposition lncRNAs with respect to the relevant protein-coding region.

The Developmental Genome Anatomy Project (DGAP) (Higgins et al. 2008) and similar projects have historically explored balanced chromosomal rearrangements to establish possible relationships between genotypes and phenotypes through identifying nucleotide-level breakpoints via Sanger sequencing. Individuals with such rearrangements represent natural gene disruptions and dysregulations, and their chromosomal rearrangements can serve as ideal signposts for annotating the human genome. Unlike for protein-coding genes, it is hard to predict the pathogenicity of lncRNA variants because they are not translated and consequently frameshift and nonsense mutations may not disrupt their function. Here, we employ a foundational approach in human genetics using chromosomal rearrangements to interrogate potential phenotypic impacts of disrupted lncRNAs and their genomic repositioning resulting in dysregulation. Both disruption and dysregulation of lncRNAs therefore may increase the diagnostic yield of developmental disorders. We venture to make a call out to cytogeneticists to employ further the power of chromosomal rearrangements in yet another opportunity to contribute to annotating the genome, recognizing that there are many patients and families who still await diagnoses.

## Methods

### Human Subjects

Study ID numbers are a consecutive alphanumeric list that are not known outside of the research group. The Partners HealthCare System Internal Review Board (IRB) gave ethical approval for this work under protocol number 1999P003090.

### Breakpoint Mapping

Genomic DNA from DGAP probands was sequenced to identify chromosomal breakpoints at nucleotide-level according to the previously published protocol (Talkowski et al. 2011; Hanscom and Talkowski 2014). Sanger sequencing results were aligned to the human genome using the UCSC Genome Browser BLAT tool (Kent 2002). Breakpoints were also compiled from previous publications (Talkowski et al. 2012b; Redin et al. 2017; Lowther et al. 2022). Breakpoint positions were converted from earlier genome builds to hg38 using the UCSC Genome Browser LiftOver tool (Kent et al. 2002).

### Additional Genetic Analyses

To ensure that the phenotypes found in DGAP103 and DGAP353 could not be attributed to other variants aside from the chromosomal rearrangements, whole exome sequencing was performed for these cases by the Genomics Platform at the Broad Institute of MIT and Harvard (Cambridge, MA). Sequencing libraries were prepared from sample DNA (250 ng input) using the Twist Bioscience exome (∼35Mb target) assay (San Francisco, CA), which were then sequenced (150 bp paired end) on the Illumina NovaSeq platform (Illumina, San Diego, CA) to generate a coverage of >85% of the target region at 20X read depth or greater. Sequencing data for each of the samples was processed through an internal pipeline using the BWA aligner for mapping to the human genome (GRCh38/hg38) and variant calling was performed using the Genome Analysis Toolkit (GATK) HaplotypeCaller package. Variants were then annotated and assessed for pathogenicity using the Seqr software (Pais et al. 2022). Variants with >1% MAF were filtered out and variants in genes with an association with disease were prioritized for analysis. No candidate variants associated with either phenotype were found. For DGAP355, whole exome sequencing (GeneDX XomeDxXpress) was negative for relevant variants. For DGAP148, array comparative genomic hybridization was previously performed and found to be normal (Redin et al. 2017).

### TAD Analysis and Visualization of Chromatin Interactions

TAD boundary positions previously identified by Dixon and colleagues (Dixon et al. 2012) were converted from hg18 to hg38 using the UCSC Genome Browser LiftOver tool (Kent et al. 2002). BEDTools was used to identify TADs that included DGAP breakpoints (Quinlan and Hall 2010). The USCS Genome Browser (Kent et al. 2002) was used to display these TAD regions. Along with the genes within these regions, we also displayed chromatin interactions identified through micro-C studies from H1-hESCs (Krietenstein et al. 2020).

### Temporal Bone Computerized Tomography (CT)

Axial temporal bone CT without contrast for the mother of DGAP353 consisted of helical images with the following parameters: Discovery STE system (General Electric Healthcare, Waukesha, WI); 0.625 mm slice thickness, effective mAs of 17, mA of 158, rotation time of 2161 milliseconds, pitch of 0.5625 and kvp of 140. Images were viewed in a plane parallel to that of the horizontal semicircular canal. Coronal reformatted images were obtained in a plane perpendicular to the axial images at 0.74 mm thickness.

Axial temporal bone CT without contrast for DGAP353 consisted of helical images with the following parameters: Discovery STE system (General Electric Healthcare, Waukesha, WI); 0.625 mm slice thickness, effective mAs of 27, mA of 246, rotation time of 2161 milliseconds, pitch of 0.5625 and kvp of 100. Images were viewed in a plane parallel to that of the horizontal semicircular canal. Coronal reformatted images were obtained in a plane perpendicular to the axial images at 0.70 mm thickness.

## Results

### Identification of Human Subjects with Disrupted lncRNAs

We evaluated 279 cases of balanced chromosomal abnormalities and selected 191 cases with resolved breakpoints indicating a “simple” rearrangement (*i.e*., breakpoints at only two genomic locations and no significant genomic imbalance) for further analysis (**Table S1**). Using the most recent Human Gencode Reference, Release 45, GRCh38.p14 (Frankish et al. 2021), we then identified 66 cases in which at least one breakpoint overlapped a lncRNA (**Table S2 and Table S3**). Overall, 79 unique lncRNAs were directly disrupted in these cases, and four lncRNAs including *MEF2C-AS1* and *ENSG00000257522* were each disrupted in two unrelated individuals. In 30 of the cases, no genes of any other class aside from lncRNAs were directly disrupted by the breakpoints. In this report, we primarily focus on two cases (DGAP353 and DGAP103) as examples of the potential value of assessing lncRNAs as diagnostic etiologies, and six additional cases are presented for further investigation (**Table 1**).

**Table 1.** Details regarding the breakpoints and the disrupted genes for the cases highlighted in this manuscript. Genomic coordinates refer to GRCh38/hg38.

### Clinical Report of DGAP353

The proband DGAP353 was diagnosed during gestation when her healthy mother (20-24 years old) underwent amniocentesis performed following an abnormal maternal serum screen for an elevated risk for trisomy 21. An apparently balanced translocation was detected in the female fetus between the long arms of chromosomes 14 and 17. Parental chromosome analyses revealed maternal inheritance and apparent structural identity to the maternal t(14;17) rearrangement. DGAP353’s G-banded karyotype is described as 46,XX,t(14;17)(q24.3;q23)mat and the mother’s karyotype is 46,XX,t(14;17)(q24.3;q23). No clinical abnormalities were observed in the fetus and the pregnancy was continued. DGAP353 began developing signs of hearing loss between the ages of 10-14 years old, and her hearing loss was found to be primarily sensorineural with a conductive element. Around this time, surgery was performed to rectify the conductive abnormalities, but the sensorineural hearing loss remained. The mother of DGAP353 began wearing hearing aids around 35-44 years of age, after a gradual decline in hearing for an unspecified time period. Both DGAP353 and her mother were otherwise healthy, typical of nonsyndromic deafness of unknown genetic etiology. Computerized tomography (CT) imaging of the temporal bones of DGAP353 and her mother revealed abnormalities such as unusually small sinus timpani and narrowing of the round and oval windows (**Fig. S1**).

### Breakpoint analysis of DGAP353

Both DGAP353 and her mother harbor a translocation between chromosomes 14 and 17 with a 7 base-pair (bp) insertion of DNA of non-templated origin at the breakpoint in the der(17) chromosome (**Fig. 1A**). Following suggested nomenclature (Ordulu et al. 2014), the next-generation cytogenetic nucleotide level research rearrangement is described in a single line as:

46,XX,t(14;17)(q24.3;q23)mat.seq[GRCh38] t(14;17)(14pter→14q23.3(+)(65,855,3{58-60}):: 17q23.2(+)(61,393,84{1-3})→17qter;17pter→17q23.2(+)(61,393,812)::TATATAC::14q23.3(+) (65,855,359)→14qter)mat.

**Figure 1.** A) Chromosome diagrams depict the translocation between 14q23.3 and 17q23.2 in DGAP353. Above, TADs containing the breakpoints are shown, with the breakpoint positions indicated by vertical yellow bars. Protein-coding genes are shown in blue and non-coding genes in green, with a single isoform depicted per gene. TAD borders were defined in (Dixon et al. 2012). Triangular contact maps display micro-C data from (Krietenstein et al. 2020). B) Expanded view of the genomic region surrounding the 14q23.3 breakpoints in DGAP353. C) Expanded view of the genomic region surrounding the 17q23.2 breakpoints in DGAP353. The directly disrupted lncRNA *TBX2-AS1* is highlighted in red. *ENSG00000267131* has been identified as an isoform of *TBX2-AS1* by LNCipedia (Volders et al. 2019).

### TBX2-AS1 is a candidate lncRNA for an association with hearing loss

The DGAP353 breakpoints do not overlap any genes on chromosome 14 (**Fig. 1B**); however, this translocation results in the direct disruption of the lncRNA *TBX2-AS1* from chromosome 17 (**Fig. 1C**). The Gencode annotation also lists the lncRNA *ENSG00000267131* as a separate gene that is disrupted by these breakpoints, however this has been identified as an isoform of *TBX2-AS1* by LNCipedia (Volders et al. 2019). While little is known regarding the biological role of *TBX2-AS1*, particularly in the context of hearing, the orthologous mouse lncRNA (*2610027K06Rik*) has been detected in the cochlear and vestibular sensory epithelium of embryonic and postnatal mice (identified as “XLOC_007930”) (Ushakov et al. 2017). Using the Gene Expression Analysis Resource (gEAR) portal (Orvis et al. 2021), we further found that while *Tbx2-as1* is detected in supporting cell types (pillar and Deiters cells), it is predominantly expressed by sensory inner hair cells, as determined through cell-type-specific RNA-seq (Liu et al. 2018). Thus, the expression pattern of *Tbx2-as1* is consistent with the finding that the hearing loss demonstrated by DGAP353 and her mother is primarily sensorineural.

The lncRNA gene *TBX2-AS1* exists in a divergent configuration with the protein-coding gene *TBX2*. Divergent lncRNAs are a particularly interesting class because they have often been found to regulate their neighboring gene. In many cases, the divergent lncRNA affects expression of the neighbor in *cis* (Luo et al. 2016; Wang et al. 2020). Some divergent lncRNAs have also been found to modulate the downstream functions of the protein generated by the neighboring gene. For example, the divergent lncRNA *Six3OS* neighbors the gene *SIX3* and regulates the activity of the SIX3 protein by functioning as a molecular scaffold (Rapicavoli et al. 2011). Similarly, *Paupar* is a lncRNA that is divergent to *Pax6* and physically interacts with the PAX6 protein to affect how this transcription factor regulates target genes (Vance et al. 2014; Pavlaki et al. 2018). It has recently been proposed that *TBX2-AS1* may function in a similar manner to regulate *TBX2* target genes in neuroblastoma cells (Modi et al. 2023). Thus, knowledge regarding the function of the protein-coding member of a divergent pair can provide insights into the potential biological role of the lncRNA partner.

Intriguingly, *TBX2* has previously been linked with hearing and inner ear development. In mice, *Tbx2* has been associated with otocyst patterning in inner ear morphogenesis, as mouse models in which *Tbx2* was conditionally knocked out exhibit cochlear hypoplasia (Kaiser et al. 2021). Previous studies have also shown that deletions encompassing *TBX2* and *TBX2-AS1* are found in individuals with hearing loss, albeit in conjunction with other deleted genes (Ballif et al. 2010; Nimmakayalu et al. 2011; Schönewolf-Greulich et al. 2011). In addition, a recent study has shown that *Tbx2* is required for inner hair cell and outer hair cell differentiation, demonstrating that it is a master regulator of hair cell fate (García-Añoveros et al. 2022). Therefore, we suggest that the translocation disrupting *TBX2-AS1* in DGAP353 and her mother may lead to altered expression or function of *TBX2*, ultimately resulting in the phenotype of hearing loss.

### Clinical Report of DGAP103

The proband DGAP103 was referred to DGAP between 5-9 years of age with a complex overgrowth phenotype that we previously reported (Ligon and Moore et al., 2005). Between 0-4 years of age DGAP103 exhibited premature dentition, including potential supernumerary teeth identified by panoramic dental X-ray. Around this time, additional features including height, weight, and occipital-frontal circumference were all at or above the 95^th^ percentile. Magnetic resonance imaging performed around this time due to macrocephaly revealed a two cm right cerebellar lesion ventral to cerebellar nuclei and stable by serial imaging as of 5-9 years of age. A bone marrow biopsy was performed at 5-9 years of age due to mild thrombocytopenia and leukopenia (platelet counts as low as 60,000) with trilineal hematopoiesis and mildly reduced cellularity but no evidence of malignancy. A G-banded cytogenetic analysis was performed and reported as a constitutional 46,XY,inv(12)(p12.2q15), which was later revised through the DGAP study to 46,XY,inv(12)(p11.22q14.3)dn. Upon enrollment in DGAP at 5-9 years of age, DGAP103 had extreme overgrowth (height at the 50%ile for 15-year-old males, weight at the 50%ile for 14-year-old males, and head circumference at the 50%ile for adult males), facial dysmorphism, brachydactyly of hands and feet with shortened distal phalanges and redundant curved nails, and bilateral lower extremity nodules of adipocytes and fibrovascular tissue consistent with lipomas.

### Breakpoint analysis of DGAP103

We have now performed breakpoint analysis, including Sanger sequencing confirmation, which determined that DGAP103 harbors a pericentric inversion of chromosome 12 with a 9bp insertion of DNA of non-templated origin (**Fig. 2A**). Following suggested nomenclature (Ordulu et al. 2014), the next-generation cytogenetic nucleotide level research rearrangement is described in a single line as: 46,XY,inv(12)(p12.2q15)dn.seq[GRCh38] inv(12)(pter→p11.22(+)(28,843,402)::TCTCAAAAA::q14.3(-)(65,851,667)→p11.22(-)(28,843,40{4})::q14.3(+)(65,851,66{9})→qter)dn.

**Figure 2.** A) Chromosome diagrams depict the inversion between 12p11.22 and 12q14.3 in DGAP103. Above, TADs containing the breakpoints are shown, with the breakpoint positions indicated by vertical yellow bars. Protein-coding genes are shown in blue and non-coding genes in green, with a single isoform depicted per gene. TAD borders were defined in (Dixon et al. 2012). Triangular contact maps display micro-C data from (Krietenstein et al. 2020). B) Expanded view of the genomic region surrounding the 12p11.22 breakpoints in DGAP103. C) The micro-C data for the TAD surrounding the 12p11.22 breakpoints in DGAP103 is expanded and annotated to highlight the interactions between the *PTHLH* locus and the potential regulatory region distal to the breakpoints. Below, the layered H3K4Me1 and H3K27Ac tracks show data from the Bernstein Lab at the Broad Institute. D) Expanded view of the genomic region surrounding the 12q14.3 breakpoints in DGAP103. The directly disrupted genes, protein-coding gene *HMGA2* and lncRNA *HMGA2-AS1*, are highlighted in red.

### Evaluation of genetic etiology of brachydactyly in DGAP103

The DGAP103 breakpoints at 12p11.22 do not overlap any known genes (**Fig. 2B**), but they occur within a TAD that contains *PTHLH*, a protein-coding gene. *PTHLH* is the only gene in a 5 Mb region around the breakpoint on the short arm of chromosome 12 with a pHaplo score above the threshold of 0.86 (*PTHLH* pHaplo = 0.92), indicating that it is predicted to exhibit haploinsufficiency (Collins et al. 2022). While our initial study of DGAP103 left the cause of brachydactyly unaddressed (Ligon and Moore et al., 2005), it was subsequently reported that deletions and point mutations in *PTHLH* result in brachydactyly (Klopocki et al. 2010; Bae et al. 2018; Reyes et al. 2019). Moreover, *PTHLH* is described by ClinGen as having sufficient evidence for haploinsufficiency causing brachydactyly type E2 (Rehm et al. 2015). In DGAP103, *PTHLH* is not directly disrupted by the chromosomal inversion; however, previously published data from micro-C studies to identify genome-wide chromatin interactions (Krietenstein et al. 2020) demonstrate that the *PTHLH* locus interacts with a region in 12p11.22 that is centromeric to the breakpoints in DGAP103, which would separate this region from *PTHLH*. This region exhibits chromatin modifications associated with enhancer activity such as H3K4Me1 and H3K27Ac (**Fig. 2C**), as determined by chromatin immunoprecipitation analysis from the ENCODE consortium (ENCODE Project Consortium 2012). Thus, altered regulation of *PTHLH* is the most likely genetic etiology for brachydactyly.

### Reevaluation of genetic etiology of overgrowth phenotypes in DGAP103 upon annotation of the lncRNA HMGA2-AS1

The DGAP103 breakpoints at 12q14.3 disrupt most of the isoforms of the protein-coding gene *HMGA2* (also known as *HMGI-C*) within the canonical third intron, as well as one isoform of its antisense lncRNA *HMGA2-AS1* (**Fig. 2D**). At the time of our initial study of DGAP103, *HMGA2-AS1* had not yet been identified, but *HMGA2* had been described as a member of the high-mobility group AT-hook (HMGA) family, which bind to DNA through three conserved AT-hook domains (Zhou and Chada 1998). *HMGA2* was already known to play crucial roles in the regulation of growth, with *Hmga2*-null mice exhibiting substantially reduced body size, while heterozygous mice are more mildly affected (Zhou et al. 1995). More recently, microdeletions in 12q14 that include *HMGA2* have been identified in individuals with short stature (Lynch et al. 2011; Alyaqoub et al. 2012), and disruptive variants in *HMGA2* have been found to cause fetal growth restriction (Abi Habib et al. 2018), supporting the role of *HMGA2* in human growth. Conversely, certain disruptions within *HMGA2* had been linked to increased proliferation, and chromosomal rearrangements within the canonical third intron of *HMGA2* had been described as the most frequent chromosomal aberration in human tumors (Kazmierczak et al. 1998). In some cases, chromosomal rearrangements had been identified in mesenchymal tumors including lipomas that result in the fusion of the *HMGA2* DNA binding domains to other regulatory domains (Ashar et al. 1995; Schoenmakers et al. 1995). Therefore, we previously tested whether a fusion product between the first three exons of *HMGA2* and *PTHLH* might exist in DGAP103, however no fusion product was detected (Ligon and Moore et al., 2005). It had also been demonstrated that overexpression of truncated *Hmg2a* causes gigantism and lipomatosis in transgenic mice (Battista et al. 1999). Thus, our initial study of DGAP103 proposed that truncation of *HMGA2*, through an inversion with a breakpoint in canonical intron 3 that leaves canonical exons 1-3 intact, was the most likely genetic etiology for overgrowth and multiple lipomas (Ligon and Moore et al., 2005).

Since then, continued annotation of the human transcriptome has provided substantially greater detail regarding the region surrounding the 12q14.3 breakpoints in DGAP103. The most recent annotation, Human Gencode Reference, Release 45, GRCh38.p14 (Frankish et al. 2021), now includes 10 isoforms of *HMGA2* and reveals that two short isoforms naturally terminate centromeric to the DGAP103 breakpoints (**Fig. 2D**). Unlike the longer isoforms of *HMGA2*, these two short isoforms would remain intact in DGAP103, but they would be repositioned to the short arm of the derivative chromosome 12.

Additionally, updated transcriptome annotations have uncovered the lncRNA *HMGA2-AS1* (**Fig. 2D**). This lncRNA is antisense to *HMGA2* and is completely overlapped by the *HMGA2* gene but transcribed from the opposite strand. Such antisense lncRNAs can function to modulate the expression of their overlapping partner in *cis* (Statello et al. 2021). More broadly, lncRNAs of several different classes have been found to regulate their neighboring genes in *cis* (Ferrer and Dimitrova 2024). For instance, mouse models have shown that the lncRNA *Chaserr* represses the expression of its neighbor *Chd2* in an allele-specific manner, demonstrating that it functions strictly in *cis* (Rom et al. 2019). Interestingly, *Chd2* promotes the expression of *Chaserr*, and thus these neighbors participate in a regulatory feedback loop in which the *Chaserr* lncRNA serves as a sensor to tightly maintain appropriate levels of *Chd2*. A recent pre-print manuscript has further demonstrated that human *CHASERR* similarly regulates *CHD2* in *cis*; individuals with *de novo* deletions in the *CHASERR* locus exhibit increased *CHD2* expression from the neighboring allele, leading to severe developmental delay and facial dysmorphisms (Ganesh et al. 2024). Intriguingly, it has recently been found that another member of the *HMGA* family, *HMGA1*, is repressed by the nearby lncRNA *HMGA1-lnc* (Stewart et al. 2020). *HMGA2-AS1* could similarly play a critical role in regulating *HMGA2*.

New analyses have also led to a refined understanding of *HMGA2* and its effects on proliferation. It was previously thought that either the creation of an *HMGA2* fusion product or the truncation of *HMGA2* were required to cause overgrowth phenotypes, including mesenchymal tumors (Fedele et al. 1998; Battista et al. 1999). However, it has more recently been shown that the overexpression of either full length or truncated human *HMGA2* in differentiated mesenchymal cells is sufficient to cause mesenchymal tumors in transgenic mouse models, and it is now proposed that the overexpression of the three *HMGA2* DNA-binding domains is the key requirement for these phenotypes (Zaidi et al. 2006). Importantly, the two short isoforms of *HMGA2* contain these DNA-binding domains, with identical sequence to that from the canonical full length *HMGA2* isoform. Thus, the overgrowth phenotypes of DGAP103 could be explained by altered regulation of *HMGA2* through the repositioning of its short isoforms to a new genomic location that lacks the antisense lncRNA *HMGA2-AS1*. Indeed, we previously detected increased expression of *HMGA2* in a lymphoblastoid cell line from DGAP103 (Ligon and Moore et al., 2005). Thus, we propose that the DGAP103 chromosomal rearrangement separating the short isoforms of *HMGA2* from the antisense lncRNA *HMGA2-AS1* may lead to increased expression of *HMGA2*, resulting in the phenotypes of overgrowth and multiple lipomas.

### Recurrent disruptions of the lncRNA MEF2C-AS1 in individuals with neurological phenotypes

We additionally identified two cases, DGAP191 and DGAP218, with chromosomal rearrangements that disrupt the lncRNA *MEF2C-AS1*. Following suggested nomenclature (Ordulu et al. 2014), the next-generation cytogenetic nucleotide level research rearrangements are described in a single line as:

<u>DGAP191:</u> 46,XY,t(5;7)(q14.3;q21.3)dn.seq[GRCh38] t(5;7)(5pter→5q14.3(+)(89,411,06{3-5}) ::7q21.3(+)(94,378,2{48-50})→7qter;7pter→7q21.3(+)(94,378,25{3-5})::5q14.3(+)(89,411,07{ 0-2})→5qter)dn.

<u>DGAP218:</u> 46,XX,inv(5)(p12q13.1)dn.seq[GRCh38] inv(5)(pter→p14.2(+)(24,272,19{3})::q14.3(-)(89,105,02{6})→p14.2(-)(24,272,189)::TATTTATATGACAAG::q14.3(+)(89,105,031) →qter)dn.

In both cases, the 5q14.3 breakpoints directly disrupt the lncRNA *MEF2C-AS1*. In DGAP191, the 7q21.3 breakpoints additionally overlap the lncRNA *ENSG00000285090*, but no protein-coding genes are directly disrupted (**Fig. 3**). In DGAP218, *MEF2C-AS1* is the only gene of any class that is directly disrupted (**Fig. 4**).

**Figure 3.** A) Chromosome diagrams depict the translocation between 5q14.3 and 7q21.3 in DGAP191. Above, TADs containing the breakpoints are shown, with the breakpoint positions indicated by vertical yellow bars. Protein-coding genes are shown in blue and non-coding genes in green, with a single isoform depicted per gene. TAD borders were defined in (Dixon et al. 2012). Triangular contact maps display micro-C data from (Krietenstein et al. 2020). B) Expanded view of the genomic region surrounding the 5q14.3 breakpoints in DGAP191. The directly disrupted lncRNA *MEF2C-AS1* is highlighted in red. C) Expanded view of the genomic region surrounding the 7q21.3 breakpoints in DGAP191. The directly disrupted lncRNA *ENSG00000285090* is highlighted in red.

**Figure 4.** A) Chromosome diagrams depict the inversion between 5p14.2 and 5q14.3 in DGAP218. Above, TADs containing the breakpoints are shown, with the breakpoint positions indicated by vertical yellow bars. Protein-coding genes are shown in blue and non-coding genes in green, with a single isoform depicted per gene. TAD borders were defined in (Dixon et al. 2012). Triangular contact maps display micro-C data from (Krietenstein et al. 2020). B) Expanded view of the genomic region surrounding the 5p14.2 breakpoints in DGAP218. C) Expanded view of the genomic region surrounding the 5q14.3 breakpoints in DGAP218. The directly disrupted lncRNA *MEF2C-AS1* is highlighted in red.

We previously reported both of these individuals as part of a larger set of cases with breakpoints in 5q14.3 (Redin et al. 2017). This region is of particular interest due to 5q14.3 microdeletion syndrome, which is characterized by neurological phenotypes including intellectual disability and epilepsy (Zweier and Rauch 2012). This syndrome is now recognized to be driven by decreased *MEF2C* expression, either through direct disruption of *MEF2C* or due to distal mutations (Zweier and Rauch 2012). Indeed, when we previously described DGAP191 and DGAP218 (Redin et al. 2017), we noted that their phenotypes were similar to individuals with direct *MEF2C* disruptions. Furthermore, we determined that levels of *MEF2C* expression were reduced in lymphoblastoid cell lines from both DGAP191 and DGAP218 (Redin et al. 2017); however, no mention was made of *MEF2C-AS1*. Recent studies have further elucidated the functional effects of altering *MEF2C* or its topological organization (Mohajeri et al. 2022), but the potential role of *MEF2C-AS1* remains unclear.

While there is still little known regarding the function of *MEF2C-AS1*, it has recently been found that *MEF2C-AS1* can positively regulate the expression of *MEF2C* in human cervical cancer cell lines (Guo et al. 2022). Interestingly, *MEF2C-AS1* is transcribed through multiple putative enhancers of *MEF2C* (D’haene et al. 2019), providing a potential mechanism for this lncRNA to regulate expression of its neighboring gene, as has previously been described for lncRNAs such as *Bendr* (Engreitz et al. 2016) and *Uph* (Anderson et al. 2016). Thus, for DGAP191 and DGAP218 we now propose that the disruption of *MEF2C-AS1* leads to decreased expression of *MEF2C*, resulting in neurological phenotypes.

### The lncRNA ENSG00000257522 is recurrently disrupted in individuals with microcephaly

Our analysis further identified two cases, DGAP245 and NIJ1, with chromosomal rearrangements that disrupt the lncRNA *ENSG00000257522* (**Fig. 5 and Fig. 6**). These individuals exhibit shared phenotypes (**Table S1**) including microcephaly and defects of the corpus callosum. Following suggested nomenclature (Ordulu et al. 2014), the next-generation cytogenetic nucleotide level research rearrangements are described in a single line as:

<u>DGAP245:</u> 46,XY,t(3;14)(p23;q13)dn.seq[GRCh38] t(3:14)(3qter→3p22.2(-)(36,927,959)::CATTTGTTCAAATTTAGTTCAAATGA::14q12(+)(29,276,117)→14qter;14pter→14q12(+) (29,276,10{8-9})::3p22.2(-)(36,927,6{49-50})→3pter)dn.

<u>NIJ1:</u> 46,XX,t(8;14)(q21.2;q12)dn.seq[GRCh38] t(8;14)(8pter→8q21.12(+)(78,898,16{9})::14q12(+)(29,296,33{1})→14qter;14pter→14q12(+)(29,296,328)::AAAT::8q21.12(+) (78,898,172)→8qter)dn.

**Figure 5.** A) Chromosome diagrams depict the translocation between 3p22.2 and 14q12 in DGAP245. Above, large regions containing the breakpoints are shown, with the breakpoint positions indicated by vertical yellow bars. The region shown surrounding 14q12 is a TAD, with its borders previously defined in (Dixon et al. 2012). No TAD was defined surrounding 3p22.2, so instead the region including 1Mb on either side of the breakpoints is displayed. Protein-coding genes are shown in blue and non-coding genes in green, with a single isoform depicted per gene. Triangular contact maps display micro-C data from (Krietenstein et al. 2020). B) Expanded view of the genomic region surrounding the 3p22.2 breakpoints in DGAP245. C) Expanded view of the genomic region surrounding the 14q12 breakpoints in DGAP245. The directly disrupted lncRNAs *ENSG00000258028* and *ENSG00000257522* are highlighted in red.

**Figure 6.** A) Chromosome diagrams depict the translocation between 8q21.13 and 14q12 in NIJ1. Above, TADs containing the breakpoints are shown, with the breakpoint positions indicated by vertical yellow bars. Protein-coding genes are shown in blue and non-coding genes in green, with a single isoform depicted per gene. TAD borders were defined in (Dixon et al. 2012). Triangular contact maps display micro-C data from (Krietenstein et al. 2020). B) Expanded view of the genomic region surrounding the 8q21.13 breakpoints in NIJ1. C) Expanded view of the genomic region surrounding the 14q12 breakpoints in NIJ1. The directly disrupted lncRNAs *ENSG00000258028* and *ENSG00000257522* are highlighted in red.

In both cases, the 14q12 breakpoints directly disrupt the lncRNA *ENSG00000257522* as well as the overlapping antisense lncRNA *ENSG00000258028*. In DGAP245, the 3p22.2 breakpoints additionally disrupt the protein-coding gene *TRANK1* (**Fig. 5B**), however this gene is not predicted to be haploinsufficient (pHaplo = 0.29) (Collins et al. 2022) and it has not been implicated in any human phenotypes by OMIM. In NIJ1, the 8q21.12 breakpoints disrupt the lncRNA *MITA1* (**Fig. 6B**). Given that the only shared disruptions between these cases are to the lncRNAs *ENSG00000257522* and *ENSG00000258028*, we focused on these for further analysis.

Using the GTEx database (Lonsdale et al. 2013), we found that *ENSG00000258028* is not readily detected in neural tissue, and thus it is unlikely to cause the patient phenotypes. In contrast, *ENSG00000257522* is primarily expressed in neural tissue (**Fig. 7A**), suggesting that it could play an important neurological role. Moreover, *ENSG00000257522* exists within the same TAD as the protein-coding gene *FOXG1*, disruptions in which have been associated with a variant of Rett syndrome (MIM # 613454) (Ariani et al. 2008) as well as *FOXG1* syndrome (Kortüm et al. 2011). Core phenotypes of these syndromes include microcephaly and corpus callosum defects, implicating *FOXG1* dysregulation as the underlying genetic etiology in DGAP245 and NIJ1. Thus, we sought to identify potential regulatory elements that could be disrupted by the chromosomal rearrangements in these cases, and found three regions with prominent H3K4me1 chromatin modification (**Fig. 7B**), which is associated with enhancer activity (ENCODE Project Consortium 2012). Notably, one of these regions also exhibited H3K27Ac modification, which is also associated with enhancer activity (ENCODE Project Consortium 2012). Furthermore, these three regions each include a portion that has been demonstrated to drive reporter expression in neural tissue *in vivo* in transgenic mice (hs566, hs1539, and hs1168) (Visel et al. 2007), and thus these regions exert experimentally validated enhancer activity.

**Figure 7.** A) Expression of the lncRNA *ENSG00000257522* from the GTEx database (Lonsdale et al. 2013). B) Expression of the protein-coding gene *FOXG1* from the GTEx database (Lonsdale et al. 2013). C) Expanded view of the genomic region surrounding the 14q12 breakpoints in DGAP246, DGAP245, and NIJ1. Breakpoint positions are indicated by vertical yellow bars. The layered H3K4Me1 and H3K27Ac tracks show data from the Bernstein Lab at the Broad Institute. Genomic regions with experimentally validated enhancer activity (“VISTA enhancers”) are shown in red (Visel et al. 2007).

Strikingly, all three of these enhancers exist within the lncRNA *ENSG00000257522*. While the most distal enhancer is partially disrupted by the breakpoints in DGAP245, the other two enhancers remain in the appropriate position relative to *FOXG1*. In NIJ1, all three of the enhancers are proximal to the breakpoints and are not separated from *FOXG1*. Thus, these enhancers are not directly disrupted by the chromosomal rearrangements, and instead their activity could be impaired due to the disruption of the lncRNA in which they are embedded. Indeed, transcription of lncRNAs through enhancers is a well-documented mechanism through which lncRNAs can regulate gene expression (Statello et al. 2021). Thus, we propose that the lncRNA *ENSG00000257522* regulates the expression of *FOXG1* through its effects on the embedded enhancers.

Further supporting this, we also identified an individual with a complex *de novo* rearrangement that similarly disrupts *ENSG00000257522*. This individual, DGAP246, exhibits consistent phenotypes including microcephaly (Redin et al. 2017). The complex rearrangement in DGAP246 consists of 14 pairs of breakpoints, including eight breakpoints in 14q12. Overall, this results in the direct disruption of the lncRNA *ENSG00000257522* while leaving the two most proximal enhancer elements in their correct position relative to *FOXG1*. Taken together, these three cases implicate the lncRNA *ENSG00000257522* in the regulation of *FOXG1*. Additionally, previous studies have reported several individuals with *FOXG1* syndrome that harbor disruptions in this region, including a translocation in “Patient 1” that directly disrupts *ENSG00000257522* (Mehrjouy et al. 2018). Thus, we propose that disruptions of this lncRNA can cause phenotypes including microcephaly and defects of the corpus callosum, consistent with *FOXG1* syndrome.

### Potential regulation of KIRREL3 by its neighboring lncRNA ENSG00000255087

We previously described DGAP148 as an individual with a neurodevelopmental disorder including attention deficits and difficulty with spatial coordination (Talkowski et al. 2012b). We have recently received updated information from the referring clinical geneticist indicating that this individual is overall in good health but continues to treat attention-deficit/hyperactivity disorder (ADHD). She was not able to complete regular high school, however she is employed. While she does not live alone, she is autonomous for tasks of everyday living, including meals, laundry, exercise, and driving. She is also described as very sociable.

DGAP148 has a *de novo* translocation (**Fig. S2A**), and following suggested nomenclature (Ordulu et al. 2014), the next-generation cytogenetic nucleotide level research rearrangement is described in a single line as:

46,X,t(X;11)(p11.2;q23.3)dn.seq[GRCh38] t(X;11)(Xqter→Xp11.4(-)(39,882,592)::TCACTGT ACAG::11q24.2(+)(127,040,509)→11qter;11pter→11q24.2(+)(127,040,509)::CTC::Xp11.4(-) (39,882,591)→Xpter)dn.

The 11q24.2 breakpoints disrupt the lncRNA *ENSG00000255087* (**Fig. S2B**). No other genes are directly disrupted by this translocation (**Fig. S3A**).

At the time of our initial report of DGAP148 (Talkowski et al. 2012b), we were unaware of the lncRNA *ENSG00000255087*, which still lacks any PubMed publications. However, *ENSG00000255087* is approximately 20kb upstream of the protein-coding gene *KIRREL3*, which has been associated with neurodevelopmental phenotypes including attention deficits (Ciaccio et al. 2021; Querzani et al. 2023). We previously found that expression of *KIRREL3* was reduced in DGAP148 (Talkowski et al. 2012b), but the potential mechanism underlying this was unclear. Upon reanalyzing this case and determining that the lncRNA *ENSG00000255087* is directly disrupted, we used the GTEx database (Lonsdale et al. 2013) to assess the expression of *ENSG00000255087*. We find that *ENSG00000255087* is predominantly expressed in neural tissue (**Fig. S2C**), similar to the *KIRREL3* expression pattern (**Fig. S3B**) and consistent with a potential neurodevelopmental role. Considering that the translocation in DGAP148 directly disrupts *ENSG00000255087* and that DGAP148 exhibits decreased *KIRREL3* levels, we suggest that the lncRNA *ENSG00000255087* is a candidate for regulating expression of its neighboring gene *KIRREL3*. Furthermore, we propose that disruption of *ENSG00000255087* can thus lead to the neurodevelopmental phenotypes described for DGAP148.

### The lncRNA SOX2-OT is implicated in an individual with epilepsy and autism spectrum disorder

DGAP355 is a nonverbal individual with global developmental delay, autism spectrum disorder (ASD), seizures, and epilepsy, whose mother has a history of multiple miscarriages. This individual has a *de novo* translocation between chromosomes 3 and 9 (**Fig. S4A**), and following suggested nomenclature (Ordulu et al. 2014), the next-generation cytogenetic nucleotide level research rearrangement is described here in a single line as solved by liWGS: 46,XX,t(3;9)(q26.3;q21.1)dn.seq[GRCh38] t(3;9)(3pter→3q26.33(+)(181,488,756)::9q21.13(+) (74,713,321)→9qter;9pter→9q21.13(+)(74,712,100)::3q26.33(+)(181,489,591)→3qter)dn.

The 3q26.33 breakpoints occur within the lncRNA *SOX2-OT* (**Fig. S4B**), which is the only gene directly disrupted by this translocation (**Fig. S5A**). *SOX2-OT* consists of dozens of isoforms that together span a nearly 850kb genomic region. The protein-coding gene *SOX2* exists entirely within an intron of *SOX2-OT* and is transcribed in the same direction. *SOX2* is a transcription factor that serves as a crucial regulator of the potency and self-renewal capacity of several progenitor cell types (Arnold et al. 2011), and in particular *SOX2* is known to play important roles in neural progenitor cells (Graham et al. 2003). *SOX2-OT* exhibits a similar expression pattern to *SOX2*, with both genes primarily expressed in neural tissue (**Fig. S4C and Fig. S5B**). Recently, *SOX2-OT* has been found to affect *SOX2* expression in varying ways in different contexts (Shahryari et al. 2015; Knauss et al. 2018; Li et al. 2020; Yin et al. 2020). Thus, we propose that the translocation in DGAP355 that disrupts the lncRNA *SOX2-OT* may lead to dysregulated expression of *SOX2*, resulting in neurodevelopmental phenotypes including ASD and epilepsy.

### Several lncRNAs are directly disrupted in DGAP cases

Additional DGAP cases in which lncRNAs are directly disrupted are listed in **Table S2**. These lncRNAs warrant further consideration, particularly for cases in which no other genes are directly disrupted. Given the abundance of lncRNAs throughout the human genome, it is not rare for chromosomal rearrangements to disrupt lncRNAs, and yet this class of gene has remained largely overlooked. As updated human genome annotations continue to include new lncRNAs, it is increasingly likely to identify lncRNAs disrupted by chromosomal rearrangements. The cases described here emphasize the importance of carefully considering such disrupted lncRNAs when evaluating potential genetic etiologies underlying patient conditions.

## Discussion

By virtue of their noncoding nature, it is difficult to assess the pathogenicity of lncRNA variants based on standards for protein-coding genes. As such, we propose a novel framework to implicate lncRNAs based on chromosomal rearrangements that disrupt the lncRNA function as we illustrate in two DGAP case examples, one involving deafness and the other a complex phenotype including overgrowth, lipomas and brachydactyly.

Deafness/hard-of-hearing (DHH) represents the most prevalent form of sensory deficit in humans. Approximately 5% of the global population are affected by the condition, and it is mostly of a genetic etiology in developed countries (Azaiez et al. 2018). Genetically determined DHH can be subdivided into Mendelian inheritance and complex inheritance, with the former being further classified into syndromic or nonsyndromic forms (Sheffield and Smith 2019). Over 100 genes are associated with nonsyndromic forms and more than 400 with syndromic forms (Alford et al. 2014). There has been an increased interest in investigating the role the non-coding genome plays in hearing loss. lncRNAs *Meg3*, *Rubie* and *Gm15083/lnc83* have been associated with proper functioning and development of the inner ear (Avraham et al. 2022). In addition, genomic duplications responsible for the DFNA58 form of deafness have been found to include certain lncRNAs, although it is unreported as to whether a lncRNA(s) might be etiologic (Lezirovitz et al. 2020; Nascimento et al. 2022). Herein, we propose a divergent lncRNA disruption to cause human disease in a Mendelian fashion in a familial case of hearing loss.

We report the disruption of the lncRNA *TBX2-AS1* by a balanced chromosomal rearrangement that segregated with DHH from mother to daughter. Little is currently known about human *TBX2-AS1* other than that it is divergent to *TBX2*. It is not listed currently in OMIM, and available databases including gnomAD (Karczewski et al. 2020) and DECIPHER (Firth et al. 2009) cannot be used to determine the level of constraint in the human genome pool or the tolerance to haploinsufficiency for lncRNAs because these metrics are defined specifically for protein-coding genes. *TBX2*, however, has been linked to hearing and inner ear development, including through the identification of deletions encompassing *TBX2* and *TBX2-AS1* (among other genes) that were found in individuals with hearing loss (Ballif et al. 2010; Nimmakayalu et al. 2011; Schönewolf-Greulich et al. 2011).

In the DGAP353 proband presented herein, the breakpoint did not affect *TBX2* itself but interrupted *TBX2-AS1*. It has been shown that *TBX2* maps to the edge of a TAD and is linked to *TBX2-AS1* as a bi-directionally transcribed topological anchor point (tap)RNA (Amaral et al. 2018; Decaesteker et al. 2018). Expression levels of such lncRNAs have been found to be highly correlated with those of their nearest protein-coding genes, and this has also been observed between *TBX2* and *TBX2-AS1*, suggesting that *TBX2-AS1* and *TBX2* may be connected on a regulatory level (Wansleben et al. 2014; Decaesteker et al. 2018). Alternatively, *TBX2-AS1* could affect the function of the TBX2 protein, as has been demonstrated for other divergent lncRNAs (Rapicavoli et al. 2011; Vance et al. 2014; Pavlaki et al. 2018). Thus, we propose the lncRNA *TBX2-AS1* as a candidate for an association with hearing loss.

Although there is a discrepancy between the ages of onset of the mother’s and daughter’s hearing loss, this may be attributed to anticipation or to confounding environmental exposures. There is also the possibility that the mother’s hearing loss began at an earlier age than indicated because she reported a significant improvement in her hearing when habilitated with hearing aids, suggesting that her hearing began to decline at an earlier age. To assess fully whether *TBX2-AS1* disruption is the causal agent for their hearing loss, a mouse model where *TBX2-AS1* is knocked down while *TBX2* remains intact would be valuable. Additional cases of *TBX2-AS1* deleterious variants with hearing loss will be needed to confirm the proposed association.

We also present DGAP103, an individual with brachydactyly, overgrowth, and lipomas that we initially assessed nearly 20 years ago (Ligon and Moore et al., 2005). We have now performed additional analyses that have enabled us to refine the breakpoints in DGAP103 and to reinterpret the genetic etiology of the phenotypes. Since our initial assessment, it has been reported that haploinsufficiency of *PTHLH* can cause brachydactyly (Klopocki et al. 2010; Bae et al. 2018; Reyes et al. 2019). We have now identified a potential regulatory region that has previously been demonstrated to interact with the *PTHLH* genomic region through micro-C analyses (Krietenstein et al. 2020). This region is separated from *PTHLH* by the inversion in DGAP103, and thus we propose dysregulation of *PTHLH* as the genetic etiology of brachydactyly in DGAP103.

Furthermore, the inversion in DGAP103 results in the repositioning of two short isoforms of *HMGA2* away from the antisense lncRNA *HMGA2-AS1*. During our initial assessment of DGAP103, these short isoforms of *HMGA2* had not been described, and the lncRNA *HMGA2-AS1* had not yet been identified. However, we had found that the expression of *HMGA2* was increased in lymphoblastoid cells from DGAP103 (Ligon and Moore et al., 2005). This is consistent with the finding that overgrowth phenotypes including gigantism and lipomatosis can be caused by overexpression of a short version of *Hmg2a* in transgenic mice (Battista et al. 1999). Given that a prominent mechanism of lncRNA function is through the regulation of neighboring genes (Statello et al. 2021), we now propose that *HMGA2-AS1* may repress *HMGA2* expression in *cis*; due to the inversion in DGAP103, such regulation has been lost, leading to increased levels of *HMGA2* resulting in the phenotypes of overgrowth and lipomas. This showcases the need to take into account the presence of lncRNAs to provide a more complete understanding of genetic etiologies. We present DGAP103 as an example to highlight the importance of reevaluating diagnoses in view of lncRNAs which were not previously annotated upon initial evaluation.

In conclusion, we have provided examples implicating lncRNAs in hearing loss (*TBX2-AS1*) and in a complex phenotype of overgrowth, lipomas and brachydactyly (*HMGA2-AS1*). We have also identified several additional lncRNAs that warrant further investigation, including *MEF2C-AS1*, *ENSG00000257522*, *ENSG00000255087*, and *SOX2-OT*. The potential connections between these lncRNAs and patient phenotypes were uncovered due to balanced chromosomal rearrangements in these loci; as such, we propose that such rearrangements are an untapped resource to functionally annotate lncRNAs. We propose that geneticists pay special attention to potential dysregulation of lncRNAs in patients where balanced chromosomal rearrangements do not disrupt protein-coding genes in a manner consistent with the observed phenotypes. With an increasing number of chromosomal rearrangements mapped due to inexpensive whole genome sequencing and optical genome mapping, additional lncRNAs that underly developmental diseases await characterization.

## Data Availability

All data produced in the present study are available upon reasonable request to the authors.

## Acknowledgements

We thank the DGAP individuals, their families and their clinicians for participation in DGAP, and the Harvard Medical School-affiliated DGAP PIs and members of their laboratories. This study was supported by the National Institute of General Medical Sciences T32 GM007748 (awarded to C.C.M. and funding provided to R.E.A) and P01 GM061354 (awarded to C.C.M). C.C.M. is also supported by the NIHR Manchester Biomedical Research Centre. C.A.W. is an Investigator of the Howard Hughes Medical Institute and is supported by the National Institute of Neurological Disorders and Stroke 5R01NS035129 and by the Allen Discovery Center for Human Brain Evolution through the Paul G. Allen Frontiers Program. E.L.M. is supported by the National Institute on Aging K99 fellowship K99AG081456.

## Supplemental figure and table legends

**Figure S1.** A) Axial CT images of the right and left temporal bones of a 40-45 year old female, the mother of DGAP353. While the inferior basal turns appeared normal (not shown), the upper basal and middle turns of each cochlea appear flattened (wide arrow in image 1). The round windows appear mildly narrow. The right cochlear aperture (short line in image 3) measured 1.5 mm TR and the left measured 1.3 mm TR. There is variant anatomy of the internal auditory canals which appear mildly flared on axial images at the level of the porus acusticus, however normal in the coronal plane. The sinus tympani (posteromedial recess of the tympanic cavity) is unusually small bilaterally (short arrow in image 3). B) Reformatted coronal CT images of the right and left temporal bones of the mother of DGAP353. The right oval window is mildly narrow in height (long arrow in image 1). The round window is also narrowed (short arrow in image 2). Note the mildly small upper cochlear turns (arrowhead in image 3). The left oval window is also mildly narrowed and opacified. The inferior osseous margin of the tympanic segment of the facial nerve canal is not clearly seen, raising concern for dehiscence at the level of the stenotic oval window (arrow in image 4). The subjacent round window appears normal in the coronal plane. C) Axial CT images of the temporal bones of a 10-14 year old female, DGAP353. Imaging reveals normal upper cochlear turns. The cochlear aperture measures 1.6 mm on each side. The sinus tympani is unusually small bilaterally (short arrows). Note the normal right stapedial crura (long arrow in image 1). The left stapedial crura are closely approximated and indistinct (long arrow in image 2). D) Reformatted coronal images of the temporal bones of DGAP353. These images reveal that the left oval window (arrow in image 1) and round window (arrow in image 2) are stenotic. The right sided oval and round windows were also slightly narrow (not shown).

**Figure S2.** A) Chromosome diagrams depict the translocation between Xp11.4 and 11q24.2 in DGAP148. Above, TADs containing the breakpoints are shown, with the breakpoint positions indicated by vertical yellow bars. Protein-coding genes are shown in blue and non-coding genes in green, with a single isoform depicted per gene. TAD borders were defined in (Dixon et al. 2012). Triangular contact maps display micro-C data from (Krietenstein et al. 2020). B) Expanded view of the genomic region surrounding the 11q24.2 breakpoints in DGAP148. The directly disrupted lncRNA *ENSG00000255087* is highlighted in red. C) Expression of the lncRNA *ENSG00000255087* from the GTEx database (Lonsdale et al. 2013).

**Figure S3.** A) Expanded view of the genomic region surrounding the Xp11.4 breakpoints in DGAP148. B) Expression of the protein-coding gene *KIRREL3* from the GTEx database (Lonsdale et al. 2013).

**Figure S4.** A) Chromosome diagrams depict the translocation between 3q26.33 and 9q21.13 in DGAP355. Above, TADs containing the breakpoints are shown, with the breakpoint positions indicated by vertical yellow bars. Protein-coding genes are shown in blue and non-coding genes in green, with a single isoform depicted per gene. TAD borders were defined in (Dixon et al. 2012). Triangular contact maps display micro-C data from (Krietenstein et al. 2020). B) Expanded view of the genomic region surrounding the 3q26.33 breakpoints in DGAP355. The directly disrupted lncRNA *SOX2-OT* is highlighted in red. C) Expression of the lncRNA *SOX2-OT* from the GTEx database (Lonsdale et al. 2013).

**Figure S5.** A) Expanded view of the genomic region surrounding the 9q21.13 breakpoints in DGAP355. B) Expression of the protein-coding gene *SOX2* from the GTEx database (Lonsdale et al. 2013).

**Table S1.** Genetic and phenotypic details for all cases analyzed as part of this study. Genomic coordinates refer to GRCh38/hg38. Derivative A and Derivative B represent the chromosomal breakpoints listed in the order recommended by Orulu et al. 2014.

**Table S2.** Details regarding the breakpoints and the directly disrupted genes for all 66 cases in which we identified a disrupted lncRNA. The first tab lists cases in which only lncRNAs are directly disrupted. The second tab lists cases in which lncRNAs are directly disrupted along with other genes. Genomic coordinates refer to GRCh38/hg38. The disruption of the lncRNA *RMST* in DGAP032 was previously reported in (Stamou et al. 2020). The disruption of the lncRNA *LINC00299* in DGAP162 was previously reported in (Talkowski et al. 2012a).

**Table S3.** Additional details for the cases in which only lncRNAs were directly disrupted. The first tab lists the nearest protein-coding gene to each disrupted lncRNA. The second tab lists all genes of any class within 100kb of the breakpoints, excluding the lncRNAs that are directly disrupted (see Table S2 for directly disrupted lncRNAs). Genomic coordinates refer to GRCh38/hg38.

